# Chronic complications among patients with type 2 diabetes in Southern Ethiopia

**DOI:** 10.1101/2024.12.23.24319561

**Authors:** Biniyam Demisse Andarge, Desta Haftu Hayelom, Sayih Mehari Degualem, Habtamu Esubalew Bezie, Habtamu Wondmagegn, Yohannes Habtegiorgis, Muluken Bekele Sorrie, Yilma Chisha

## Abstract

**Background:** Type 2 diabetes mellitus (T2DM) is a global health concern associated with complications that significantly impact patients’ quality of life and place significant burdens on healthcare systems. While the prevalence of T2DM is rising in Ethiopia, the scope and factors contributing to its complications remain understudied. Hence, this study aimed to assess the burden and identify associated factors of chronic complications among Type 2 diabetes mellitus patients attending Wolaita Sodo University Comprehensive Specialized Hospital in Southern Ethiopia.

**Methods:** A facility-based cross-sectional quantitative study was conducted from July 1 to August 30, 2024, involving 404 systematically sampled T2DM patients. Data on sociodemographic characteristics, clinical profiles, self-care practices, and chronic complications were collected through structured interviews and medical record reviews. Descriptive statistics summarized patient characteristics, while multivariable logistic regression identified factors associated with chronic complications. Results were reported as Adjusted Odds Ratios (AORs) with 95% Confidence Intervals (CIs), and a p-value < 0.05 was considered statistically significant.

**Results:** A total of 404 type 2 Diabetes mellitus patients participated in the study, with a response rate of 97.58%. The mean age of participants was 44.80 ± 14.10 years, 41.09% had diabetes for more than 5 years, and 64.85% had suboptimal glycemic control. Among participants, 45.54% (95% CI: 40.61 - 50.54) had at least one chronic complication, and one in five had multimorbidity. The most common microvascular complications were peripheral neuropathy (14.85%) and nephropathy (9.65%), while macrovascular complications included congestive heart failure (14.11%) and cerebrovascular disorders (11.39%). Multivariable logistic regression identified older age (AOR = 2.74; 95% CI: 1.73, 4.37; p=0.002), female sex (AOR = 2.14; 95% CI: 1.12, 4.76; p=0.018), longer diabetes duration (AOR = 2.98; 95% CI: 1.41, 6.54; p<0.001), poor glycemic control (AOR = 2.02; 95% CI: 1.33, 3.09; p=0.032), hypertriglyceridemia (AOR = 2.00; 95% CI: 1.06, 3.80; p=0.001), high salt intake (AOR = 1.57; 95% CI: 1.06, 2.32; p=0.005), and physical inactivity (AOR = 1.75; 95% CI: 1.16, 2.64; p=0.025) as significant factors associated with chronic complications.

**Conclusion:** The study identified that nearly half of the T2DM patients experienced chronic complications, highlighting a need for improved prevention strategies. Targeted screening, regular monitoring of blood glucose and lipids, personalized counseling on diet and physical activity, and integrated chronic disease management should be prioritized to reduce the complications and improve health outcomes.

## Introduction

Diabetes mellitus (DM) is a chronic, heterogeneous metabolic disorder characterized by elevated blood glucose levels due to impaired insulin secretion, action, or both. Type 2 diabetes mellitus (T2DM), the most prevalent form of DM, primarily affects middle-aged and older adults (1,2). In addition to genetic predisposition, it is primarily associated with modifiable lifestyle factors, such as poor dietary habits, obesity, and physical inactivity (3)

Non-communicable diseases (NCDs) account for three-quarters of all deaths worldwide, and diabetes continues to be a major public health challenge (4). According to the International Diabetes Federation (IDF) report, an estimated 589 million people were living with diabetes worldwide, with T2DM accounting for 90% of cases, and this prevalence is projected to rise to 853 million by 2050 (5). Diabetes was responsible for approximately 3.4 million deaths and USD 1 trillion in health-related expenditures, representing 12% of global health expenditure (5,6). Sub-Saharan Africa is witnessing a rapid increase in diabetes prevalence, with an estimated 24 million people affected, and projected to reach 60 million by 2050 (5,7). In Ethiopia, the national prevalence of diabetes among the adult population is 4.4% (5). T2DM is estimated to range from 2.0% to 6.5%, and complications are a growing concern (8).

Chronic hyperglycemia in diabetes substantially raises the risk of both microvascular (retinopathy, nephropathy, neuropathy) and macrovascular complications (cardiovascular disease, stroke) (9), which can lead to severe outcomes like blindness, kidney failure, and amputations (10,11). Half of diabetes patients are unaware of their disease, and complications are more likely in patients with poor glucose control, whereas improved management can slow disease progression (12,13). Cardiovascular disease is the most common complication, with two-thirds of diabetes patients dying from myocardial infarction or stroke (14). Additional complications include end-stage renal disease (ESRD), neuropathy, and lower-extremity amputations (15,16).

The pathogenesis of diabetes complications involves multiple risk factors, some modifiable (e.g., diet, smoking, physical inactivity) and others non-modifiable (e.g., genetics) (17,18). A high BMI is a primary risk factor globally, with other significant contributors including dietary risks, tobacco use, and low physical activity (19). In response, the World Health Organization (WHO) and the United Nations have developed global action plans and Sustainable Development Goals (SDGs) to reduce the NCD burden, aiming to decrease premature mortality from NCDs by one-third by 2030 (20,21). As a WHO member, Ethiopia has adopted these frameworks and implemented a National Strategic Plan to address NCDs. However, despite these efforts, the national burden of diabetes and its complications continues to rise (22,23).

In Ethiopia, diabetes care remains challenged by limited resources, and the complexity of diabetes complications poses substantial obstacles to improving patient outcomes. As diabetic complications often go undetected until advanced stages, there is a need for early detection and proactive management strategies in clinical settings, especially in regions with limited diagnostic and treatment capacity. Addressing this need, this study explores the scope and nature of complications among patients with T2DM to improve understanding of the condition and to inform healthcare practices and policies. By investigating the factors associated with diabetes complications, this study aims to provide healthcare providers and policymakers with actionable insights that support the development of locally adapted interventions to reduce complications, improve patient quality of life, and alleviate the healthcare burden in low-resource settings.

## Materials and methods

### Study setting and period

This study was conducted at Wolaita Sodo University Comprehensive Specialized Hospital, located in Sodo Town, Wolaita Zone, Southern Ethiopia, from July 1 to August 30, 2024. Sodo Town lies approximately 329 km south of Addis Ababa and serves as the administrative center of the South Ethiopia Regional State. The hospital is the largest healthcare facility in the region, with a capacity of 370 beds distributed across four main departments: medical, gynecology/obstetrics, surgical, and pediatrics. It is staffed by 975 health professionals and 760 administrative personnel, collectively serving a catchment population of over 5 million people. The hospital has a chronic care unit that manages over 1,350 diabetic patients, 986 of whom are diagnosed with Type 2 diabetes mellitus (T2DM).

### Study design

We employed a facility-based cross-sectional study design.

### Population and eligibility criteria

We included patients with Type 2 diabetes mellitus (T2DM) attending routine follow-up at Wolaita Sodo University Comprehensive Specialized Hospital during the study period. Eligible participants were adult patients aged 18 years and older with a clinical diagnosis of T2DM, who were willing to provide information and had attended at least three follow-up visits at the hospital. We excluded patients with incomplete medical records, missing essential data such as laboratory results, or undocumented information on diabetes-related complications. Those diagnosed with Type 1 or gestational diabetes were also excluded.

### Sample size determination

The required sample size was calculated with a 43% estimated proportion of complications among diabetic patients (24), a 5% margin of error, and a 95% confidence level, yielding a sample size of 376 patients. Using these parameters, the sample size (n) was determined with the following formula:

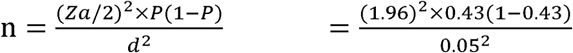

Accounting for a 10% non-response rate, the final sample size was 414 patients. Further analysis validated the adequacy of the sample size.

### Sampling technique and procedures

We used a systematic random sampling method to select participants from T2DM patients attending routine follow-up visits. Before sampling, the total list of eligible patients receiving care at the diabetic clinic during the study period was obtained. The sampling interval (k≈2) was determined by dividing the total number of eligible patients (986) by the desired sample size of 414. During the study period, the research team approached patients who visited the clinic, explained the study purpose, and obtained informed consent. Participants who agreed to participate had their clinical records reviewed, and data were collected accordingly.

### Data collection tools and procedures

#### Data collection instruments

We used a structured, pre-tested questionnaire and a chart review checklist, developed through literature review and from validated tools (24,25). The instruments were designed to capture information on socio-demographic characteristics, clinical profiles, self-care practices, and diabetes-related complications. Sociodemographic characteristics included age, sex, marital status, residence, educational level, occupation, income, and family history of diabetes. Clinical characteristics focused on duration of diabetes, body mass index (BMI), blood pressure status, glycemic control (fasting blood sugar or HbA1c), current medications, and biochemical indicators such as lipid profiles. Self-care practices assessed physical activity, dietary salt intake, foot care routines, glucometer use, smoking, and alcohol consumption. Complications are categorized as microvascular (peripheral neuropathy, nephropathy, retinopathy) or macrovascular (angina, myocardial infarction, stroke, transient ischemic attack, heart failure, and lower-extremity complications), assessed through medical chart review and/or patient self-report, corroborated with clinical documentation.

We initially prepared the questionnaire in English, translated it into Amharic and Wolaytta with the help of bilingual experts, and then back-translated it into English to ensure consistency. We conducted a pilot study with 5% of the sample at a nearby health facility to refine the tool for clarity and cultural relevance.

#### Diagnosis of chronic complications

The diagnosis of complications was made by medical specialists following Ethiopian national guidelines and World Health Organization (WHO) recommendations (26,27).

##### Peripheral neuropathy

Identified through documentation of characteristic symptoms (e.g., burning, tingling, numbness) and signs (e.g., reduced sensation, absent ankle reflexes), assessed using standard foot examinations or monofilament testing.

##### Diabetic nephropathy

Diagnosed based on proteinuria, confirmed in two or more urine tests taken 1–3 months apart, and/or estimated glomerular filtration rate (eGFR) of <60 mL/min/1.73 m².

##### Diabetic retinopathy

Confirmed through fundoscopic examination, conducted by ophthalmologists using ophthalmoscopy, as documented in the patient’s medical records.

##### Macrovascular complications

coronary artery disease (angina, myocardial infarction), heart failure, and cerebrovascular events (stroke, TIA) were identified based on physician-documented diagnoses, supported by prior history, or clinical evaluation.

##### Lower-extremity complications

foot ulcers, deformities, or amputations were confirmed from documentation and/or direct observation.

All diagnoses were based on the most recent evaluations documented in patient records during follow-up visits. Where necessary, treating physicians verified the presence of complications for consistency and accuracy.

#### Data collection process and personnel

Two trained and experienced BSc nurses collected data using face-to-face interviews in private settings. Written informed consent was obtained from all participants before data collection. Medical records were reviewed immediately after the interviews to gather additional clinical data and verify documented complications.

The process was supervised daily by the principal investigator and a medical doctor to ensure data integrity, completeness, and adherence to protocol. All completed questionnaires and checklists were reviewed at the end of each day, and any discrepancies were promptly clarified using patient records or direct follow-up with the participants.

### Operational definitions

#### Chronic complications of diabetes mellitus

Chronic complications are defined as the presence of one or more vascular complications in a diabetic patient, as diagnosed by a physician. These complications are classified as either microvascular or macrovascular. Microvascular complications include neuropathy, nephropathy, and retinopathy. Macrovascular complications include coronary artery disease (angina and myocardial infarction), cerebrovascular disorders (stroke and transient ischemic attack), congestive heart failure, and peripheral artery disease (foot ulcers and leg amputations). Patients without any documented chronic complications are categorized as having no complications (28).

#### Multimorbidity

Defined as the presence of two or more diabetes-related complications in a single individual as documented in medical records.

#### Glycemic status

was determined by averaging three consecutive fasting blood sugar measurements and/or the most recent HbA1c. Control was classified as: optimal (FBS 80– 130⍰mg/dL or HbA1c <⍰7%), and suboptimal (FBS <⍰70 or >⍰130⍰mg/dL, or HbA1c ≥⍰7%) (29).

#### Blood pressure category

Blood pressure measurements were taken from three observations, and the average of the second and third readings was recorded for analysis. Raised BP was defined as systolic blood pressure (SBP) ≥ 140 mmHg and/or diastolic blood pressure (DBP) ≥ 90 mmHg (30).

#### Anthropometric measurement

Body mass index (BMI) is calculated by dividing the participant’s weight (kg) by height squared (m²) after data collection. Classified as Normal: BMI of 18.5-24.9 kg/m², Overweight: BMI of 25-29.9 kg/m², and Obese: BMI of ≥30 kg/m².

#### Dietary salt intake

self-reported average daily salt consumption based on a 24-hour dietary recall. Participants consuming ≤ 2300 mg/day (≤ 1 teaspoon/day) were classified as having low to moderate salt intake, while those consuming > 2300 mg/day (> 1 teaspoon/day) were categorized as having high salt intake (31).

#### Physical activity

self-reported total minutes spent per week in moderate-to-vigorous physical activity. Participants who engaged in at least 150 minutes of moderate-intensity aerobic activity per week or 75 minutes of vigorous-intensity activity per week were classified as physically active, while those who did not meet these criteria were considered inactive (31).

#### Alcohol consumption

was measured based on self-reported daily intake. Women consuming more than one drink per day and men consuming more than two drinks per day were categorized as having high alcohol consumption (31).

#### Foot care practice

Assessed through self-report using a simplified checklist based on IDF guidelines. Practices included daily inspection of feet, proper hygiene, wearing appropriate footwear, and avoiding walking barefoot. Participants practicing at least three of the four behaviors were classified as having good foot care; otherwise, poor foot care (32).

#### Social support

was measured using an eightl7litem adapted MOSl7lSSS, covering emotional, instrumental, and informational support. Responses were averaged and scaled (0–100). Participants scoring in the lowest tertile were classified as ‘low’ support, those in the middle tertile as ‘moderate,’ and the highest tertile as ‘high’ (33).

### Statistical analysis

The collected data were entered into EpiData Manager version 4.2 and exported to R software version 4.4.1 for analysis. Descriptive statistics, including frequencies, percentages, means, and standard deviations, were used to summarize the socio-demographic, clinical, and self-care characteristics of the study participants.

Bivariate logistic regression was performed to examine the crude association between each independent variable and the presence of chronic complications among Type 2 diabetes mellitus patients. Variables with a p-value < 0.25 in the bivariate analysis were considered for multivariable analysis. In addition, variables were selected for multivariable logistic regression based on theoretical relevance, prior evidence from the literature, and potential for confounding. Specifically, age, sex, diabetes duration, glycemic control, and blood pressure were retained regardless of bivariate p-values due to their established roles in complication risk.

Multivariable logistic regression was then performed to identify independent factors associated with chronic complications. Adjusted odds ratios (AORs) with 95% confidence intervals (CIs) were computed, and statistical significance was declared at a p-value < 0.05. The model’s goodness-of-fit was assessed using the Hosmer–Lemeshow test (p = 0.68), and multicollinearity was checked using the Variance Inflation Factor (VIF<5).

### Data quality assurance

We conducted a one-day training for data collectors and supervisors covering study objectives, data collection instruments, ethical guidelines, and medical record review procedures. A pre-test involving 5% of the sample size was conducted at Dilfana Primary Hospital to evaluate the data collection tools and procedures. During data collection, the principal investigator and supervisors maintained continuous oversight, conducting daily spot-checks of completed questionnaires to promptly address discrepancies. Forms were reviewed each day to verify completeness and accuracy, and double data entry was performed on a subset of questionnaires to identify and resolve any inconsistencies. Finally, biostatisticians and subject matter experts reviewed the statistical analysis to validate the study findings.

### Ethical consideration

We obtained ethical approval from the Institutional Review Board of Arba Minch University College of Medicine and Health Sciences (IRB/23166/2024) and informed consent from each participant after a clear explanation of the study objectives. For illiterate participants, trained data collectors read the information sheet and consent form aloud in their preferred language, and verbal consent was documented with a witness present. Confidentiality was maintained by omitting personal identifiers from the questionnaire, and all data were securely stored and accessible only to the research team.

## Results

### Sociodemographic factors

The study included a total of 404 diabetes mellitus patients with a response rate of 97.58%. The mean age of respondents was 44.80 ± 14.10 years, where 65.35% were above 40 years, and 57.67% were male. Most participants were married (79.21%), had at least secondary education (57.18%), and resided in urban areas (59.16%). The majority had insurance (65.59%) and reported moderate social support (53.96%). Additionally, 43.56% had a family history of diabetes (Table 1).

**Table 1:** Sociodemographic characteristics of patients with type-2 diabetes mellitus in South Ethiopia, 2024 (n=404).

| Variable | Categories | Frequency | Proportion |
| --- | --- | --- | --- |
| **Age** | Mean (SD)= 44.80 ± 14.10 |  |  |
|  | 18-39 | 140 | 34.65 |
|  | 40+ | 264 | 65.35 |
| **Sex** | Male | 233 | 57.67 |
|  | Female | 171 | 42.33 |
| **Marital status** | Married | 320 | 79.21 |
|  | Single | 77 | 19.06 |
|  | Others* | 7 | 1.73 |
| **Educational level** | No formal education | 87 | 21.53 |
|  | Primary (Grade 1–8) | 86 | 21.29 |
|  | Secondary (Grade 9–12) | 116 | 28.71 |
|  | Tertiary (12+) | 115 | 28.47 |
| **Occupation** | Paid employee | 95 | 23.51 |
|  | Merchant | 67 | 16.58 |
|  | Farmer | 63 | 15.59 |
|  | Daily laborer | 14 | 3.47 |
|  | House-wife | 85 | 21.04 |
|  | Non-employee | 17 | 4.21 |
|  | Students | 63 | 15.59 |
| **Residence** | Urban | 239 | 59.16 |
|  | Rural | 165 | 40.84 |
| **Health Insurance** | Yes | 265 | 65.59 |
|  | No | 139 | 34.41 |
| Social support | Low | 130 | 32.18 |
|  | Moderate | 218 | 53.96 |
|  | High | 56 | 13.86 |
| Family History of DM | Yes | 176 | 43.56 |
|  | No | 228 | 56.44 |

*Others\*=Widowed, Divorced*

### Clinical characteristics

The clinical characteristics of the patients revealed that 58.91% had diabetes for less than 5 years, with 46.04% on insulin monotherapy and 64.85% having suboptimal glycemic control. Additionally, 51.49% had raised blood pressure, 64.60% were either overweight or obese, 39.61% had high triglycerides (≥200 mg/dL), and 45.55% had high cholesterol levels (≥240mg/dL) (Table 2).

**Table 2:** Clinical characteristics of patients with type-2 diabetes mellitus in South Ethiopia, 2024 (n=404).

| Variable | Value | Frequency | Proportion |
| --- | --- | --- | --- |
| DM duration, years | <5 years | 238 | 58.91 |
| | $\geq 5$ years | 166 | 41.09 |
| Current regimen | Insulin monotherapy | 186 | 46.04 |
|  | Insulin + OGAs | 104 | 25.74 |
|  | OGA monotherapy | 30 | 7.43 |
|  | OGA combination therapy | 84 | 20.79 |
| Glycemic control | Optimal | 142 | 35.15 |
|  | Suboptimal | 262 | 64.85 |
| Blood Pressure | Normal | 196 | 48.51 |
|  | Raised | 208 | 51.49 |
| BMI | Normal | 143 | 35.40 |
|  | Overweight | 186 | 46.04 |
|  | Obese | 75 | 18.56 |
| Triglycerides | Normal (150–199mg/dL) | 104 | 25.74 |
|  | Borderline (150–199mg/dL) | 140 | 34.65 |
| | High ($\geq 200$mg/dL) | 160 | 39.61 |
| Total cholesterol | Normal ($< 200$mg/dL) | 105 | 25.99 |
|  | Borderline (200–239mg/dL) | 115 | 28.46 |
| | High ($\geq 240$mg/dL) | 184 | 45.55 |

### Self-care practices of patients with type 2 diabetes mellitus

Among the study participants, 58.66% did not own a glucometer, and 31.68% of participants consumed excessive salt, with 43.56% of them experiencing complications. A sedentary lifestyle was prevalent (68.32%), and those who were inactive had a higher proportion of complications. Cigarette smoking was rare (4.21%), but over half of smokers (52.94%) had complications. Alcohol consumption was high in 17.82%, with 72.22% of them developing complications (Table 3).

**Table 3:** Self-care practices of patients with type-2 diabetes mellitus in South Ethiopia, 2024 (n=404).

| Variable | Categories | Chronic complications |  |  |  |
| --- | --- | --- | --- | --- | --- |
|  |  | Yes | No | Total | Proportion |
| Having Glucometer | Yes | 55 | 112 | 167 | 41.34 |
|  | No | 114 | 123 | 237 | 58.66 |
| Dietary Salt Intake | Normal | 101 | 175 | 276 | 68.32 |
|  | High | 75 | 53 | 128 | 31.68 |
| Physical Activity | Active | 40 | 88 | 128 | 31.68 |
|  | Not Active | 158 | 118 | 276 | 68.32 |
| Foot care practice | Good | 27 | 126 | 153 | 37.87 |
|  | Poor | 183 | 68 | 251 | 62.13 |
| Cigarette smoking | No | 201 | 186 | 387 | 95.79 |
|  | Yes | 9 | 8 | 17 | 4.21 |
| Alcohol consumption | Normal | 158 | 174 | 332 | 82.18 |
|  | High | 52 | 20 | 72 | 17.82 |

### Burden and patterns of chronic complications

Among diabetes mellitus (DM) patients, 45.54% (95% CI: 40.61-50.54) developed at least one complication. Among those who developed complications, 20.05% had one, 14.60% had two, 8.42% had three, and 2.48% had developed four complications (Figure 1).

**Figure 1:**
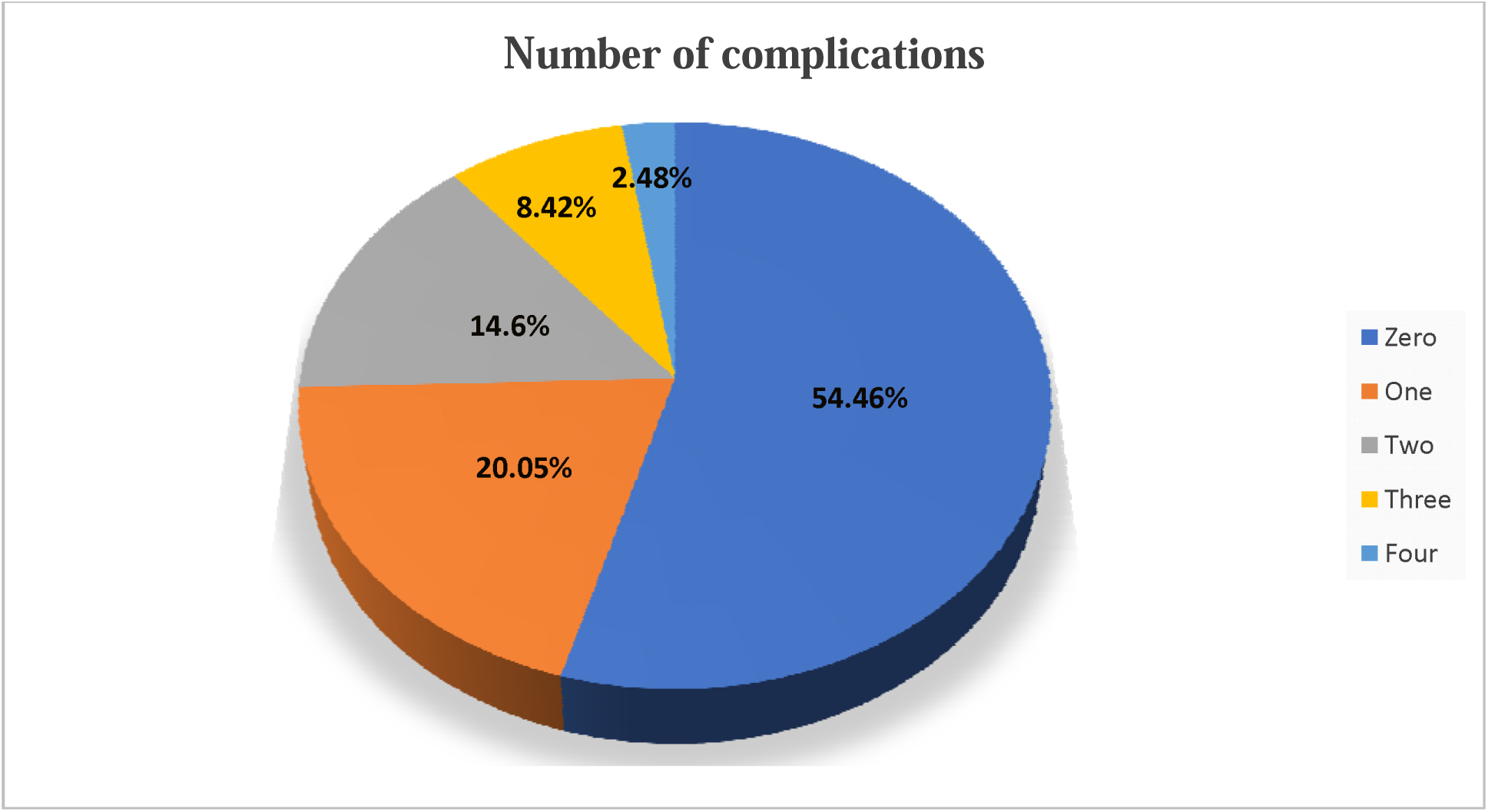
Number of complications developed among patients with type-2 diabetes mellitus in South Ethiopia, 2024. (n=404)

In this study, microvascular complications were prevalent, with peripheral neuropathy affecting 14.85% of patients, nephropathy 9.65%, and retinopathy 4.21%. In terms of macrovascular complications, congestive heart failure (CHF) affected 14.11% of patients, peripheral artery disease (lower-extremity complications) affected 9.90%, cerebrovascular disorders (stroke and transient ischemic attack) affected 11.39%, and coronary artery disease (angina or myocardial infarction) affected 4.70% of patients (Figure 2).

**Figure 2:**
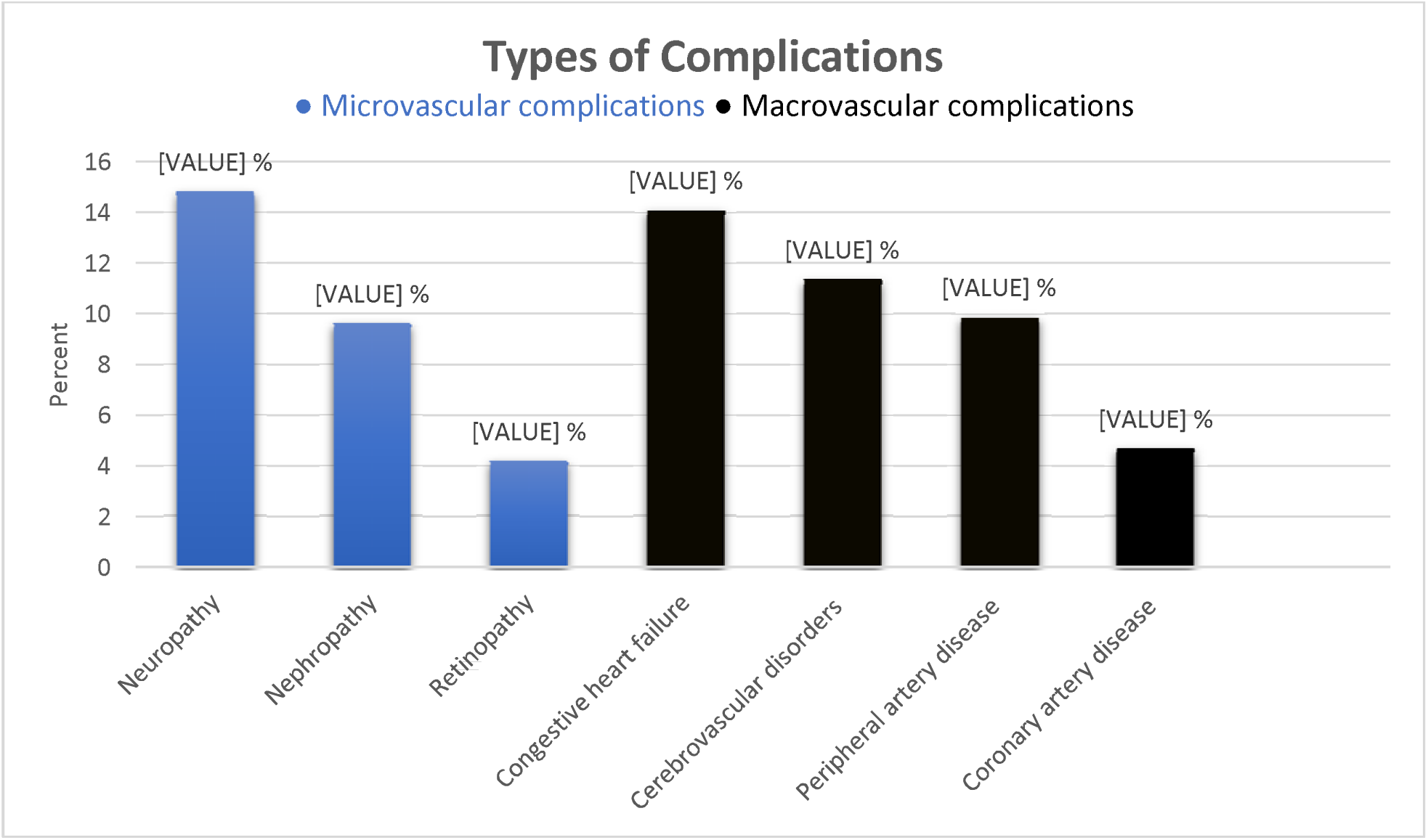
Microvascular and macrovascular complications among patients with type-2 diabetes mellitus in South Ethiopia, 2024. (n=404)

### Factors associated with chronic complications

In the bivariate logistic regression analysis, age, sex, educational level, residence, DM duration, glycemic control, blood pressure, body mass index, triglycerides, total cholesterol, salt consumption, physical activity, and glucometer use were significantly associated with chronic complications. These variables were selected for multivariable logistic regression based on a p-value < 0.25 and theoretical relevance to adjust for potential confounding.

Multivariable logistic regression analysis identified several factors significantly associated with the chronic complications among Type 2 diabetes mellitus patients. Among the sociodemographic characteristics, age and sex were significantly associated with chronic complications. Patients aged 40 years and older had nearly three times the odds of complications compared to younger patients (AOR = 2.74; 95% CI: 1.73, 4.37; p=0.002). Female patients had significantly higher odds of complications compared to males (AOR = 2.14; 95% CI: 1.12, 4.76; p=0.018).

Clinical characteristics were also independently associated with chronic complications. Patients with diabetes duration of ≥5 years had nearly threefold higher odds of complications compared to those with a shorter duration (AOR = 2.98; 95% CI: 1.41, 6.54; p<0.001). Suboptimal glycemic control was also significantly associated with complications, with affected patients having twice the odds of complications (AOR = 2.02; 95% CI: 1.33, 3.09; p=0.032). Additionally, patients with high triglyceride levels (≥200 mg/dL) had significantly greater odds of complications than those with normal levels (AOR = 2.00; 95% CI: 1.06, 3.80; p=0.001).

Among lifestyle-related variables, salt consumption and physical activity were significantly associated with chronic complications. Patients with a high dietary salt intake had 57% higher odds of chronic complications (AOR = 1.57; 95% CI: 1.06, 2.32; p=0.005). Similarly, individuals who were not physically active had 1.75 times higher odds of complications compared to those who were physically active (AOR = 1.75; 95% CI: 1.16, 2.64; p=0.025) (Table 4).

**Table 4:** Factors associated with chronic complications among patients with type-2 diabetes mellitus in South Ethiopia, 2024 (n=404).

| Variable | Categories | Chronic Complications |  |  |
| --- | --- | --- | --- | --- |
|  |  | COR (95% CI) | AOR (95% CI) | P-value |
| Age | 18-39 |  | 1 |  |
|  | 40+ | 4.46 (2.48, 8.04) | **2.74 (1.73, 4.37)\*\*** | 0.002 |
| Sex | Male |  | 1 |  |
|  | Female | 3.33 (1.15, 10.00) | **2.14 (1.12, 4.76)\*** | 0.018 |
| Educational level | No formal education |  | 1 |  |
|  | Primary (Grade 1–8) | 0.29 (0.15, 0.55) | 0.62 (0.23, 1.63) | 0.333 |
|  | Secondary (Grade 9–12) | 0.18 (0.09, 0.34) | 0.69 (0.46, 1.03) | 0.527 |
|  | Tertiary (12+) | 0.17 (0.09, 0.31) | 0.92 (0.32, 2.67) | 0.874 |
| Residence | Urban |  | 1 |  |
|  | Rural | 0.55 (0.37, 0.83) | 0.95 (0.45, 2.00) | 0.411 |
| DM duration | <5 years |  | 1 |  |
| | $\geq 5$ years | 3.42 (2.10, 5.57) | **2.98 (1.41, 6.54)\*\*\*** | 0.000 |
| **Glycemic control** | Optimal |  | 1 |  |
|  | Suboptimal | 2.68 (1.44, 4.97) | **2.02 (1.33, 3.09)\*** | 0.032 |
| **Blood Pressure** | Normal |  | 1 |  |
|  | Raised | 2.11 (1.32, 3.37) | 1.73 (0.77, 3.87) | 0.179 |
| **BMI** | Normal |  | 1 |  |
|  | Overweight | 2.76 (1.75, 4.41) | 1.35 (0.62, 2.92) | 0.148 |
|  | Obese | 3.16 (1.77, 5.70) | 1.20 (0.44, 3.32) | 0.125 |
| **Triglycerides** | Normal (<150mg/dL) |  | 1 |  |
|  | Borderline (150–199mg/dL) | 2.51 (1.19, 5.25) | 2.77 (0.77, 10.77) | 0.127 |
|  | High (≥200mg/dL) | 2.25 (1.06, 4.78) | **2.00 (1.06, 3.80)\*\*** | 0.001 |
| **Total Cholesterol** | Normal (<200mg/dL) |  | 1 |  |
|  | Borderline (200–239mg/dL) | 1.90 (0.79, 4.53) | 1.36 (0.51, 3.70) | 0.129 |
|  | High (≥240mg/dL) | 2.42 (1.04, 5.64) | 1.35 (0.31, 5.69) | 0.186 |
| **Salt Intake** | Normal |  | 1 |  |
|  | High | 2.50 (1.63, 3.86) | **1.57 (1.06, 2.32)\*\*** | 0.005 |
| **Physical Activity** | Active |  | 1 |  |
|  | Not Active | 3.01 (1.04, 8.73) | **1.75 (1.16, 2.64)\*** | 0.025 |
| **Glucometer Use** | Yes |  | 1 |  |
|  | No | 1.97 (1.30, 2.98) | 1.64 (0.71, 3.78) | 0.241 |

\* $p < 0.05$ , \*\* $p < 0.01$ , \*\*\* $p < 0.001$ ; 1=reference

## Discussion

### Burden and patterns of diabetes complications

Our study identified a significant burden of chronic complications among patients with type 2 diabetes mellitus (T2DM), in which 45.54% (95% CI: 40.61–50.54) developed at least one complication, and one in five presented with multimorbidity. Specifically, 20.05% of the patients developed one complication, 14.60% developed two, 8.42% developed three, and 2.48% developed four complications. These findings align with the earlier studies carried out in Ethiopia, which show the persistent diabetes complication burden and the need for effective management practices (24,34–36). However, the prevalence in this study is lower than compared in other reports in high- and middle-income countries (37–40) and may be attributed to differences in healthcare access, early diagnosis, and management practices that enhance complication detection and reporting.

Microvascular complications were prevalent with peripheral neuropathy (14.85%), nephropathy (9.65%), and retinopathy (4.21%) being the most common. These findings reflect the long-term impact of chronic hyperglycemia on nerve function, as reported in other settings (40,41). Similarly, macrovascular complications were also prevalent with congestive heart failure (14.11%), peripheral artery disease (9.90%), cerebrovascular disorders (11.39%), and coronary artery disease (4.70%). The high prevalence indicates the substantial cardiovascular burden associated with type 2 diabetes mellitus, which mainly contributes to morbidity and mortality in T2DM (15). The varied distribution of complications and the high prevalence of multimorbidity may indicate challenges and potential gaps in current diabetes management.

### Sociodemographic factors

Our findings indicate that sociodemographic factors such as age and sex are significantly associated with diabetes complications. Individuals aged 40 years and older have nearly threefold higher odds of complications, in line with prior research that diabetic complications, hospitalization, and mortality are more common (42,43). The cumulative effects of hyperglycemia, age-related vascular damage, and metabolic changes likely explain this association. However, emerging evidence shows that early-onset diabetes may accelerate complications due to prolonged hyperglycemia and rapid β-cell decline (44), a growing concern in sub-Saharan Africa as diabetes prevalence rises among younger populations (45). These findings emphasize the need for age-specific interventions, including early screening and management strategies tailored to younger and older individuals.

Sex differences were also observed, with female patients experiencing higher odds of chronic complications than males. Evidence suggests diabetes poses a greater risk of vascular disease for women, including coronary heart disease and stroke (46,47). Biological factors, such as hormone-related endothelial dysfunction and sex-specific fat distribution, may contribute to these differences (48). Additionally, women often receive delayed diagnoses and less preventive care, leading to more advanced vascular complications by the time of clinical presentation (49). Sociocultural barriers, including lower autonomy in health decision-making, lower income, and less access to health services, may further hinder timely care and treatment adherence among women in rural settings (50). These findings underscore sex-specific interventions in diabetes screening and management.

### Clinical characteristics

Among clinical characteristics, longer diabetes duration (≥5 years) and suboptimal glycemic control were associated with chronic complications. Prolonged exposure to hyperglycemia accelerates vascular damage through mechanisms such as endothelial dysfunction, oxidative stress, and the accumulation of advanced glycation end-products, contributing to both microvascular and macrovascular complications (36,37). This may indicate the importance of early diagnosis and glycemic control to delay disease progression.

High triglyceride levels (≥200 mg/dL) were also another factor significantly associated with chronic complications in this study. Lipid abnormalities promote atherosclerosis, endothelial inflammation, and plaque formation that can increase the risk of coronary artery disease and stroke among patients with T2DM (51,52). Elevated triglyceride levels promote the accumulation of small, dense low-density lipoprotein (LDL) particles, which are more readily oxidized and can penetrate arterial walls, triggering atherogenesis (53). Hypertriglyceridemia also increases levels of remnant cholesterol and very low-density lipoprotein (VLDL), both of which are strongly associated with cardiovascular risk (54). These findings highlight the importance of routine lipid monitoring, patient education, and integration of lipid-lowering therapy into diabetes management.

Although body mass index (BMI) and blood pressure were not statistically significant in the final model, their clinical relevance remains important. Obesity, especially visceral fat accumulation, contributes to insulin resistance, low-grade inflammation, and lipid abnormalities (55). Similarly, elevated blood pressure is a well-established risk factor for both nephropathy and cardiovascular disease in T2DM patients (56). The lack of statistical association in our analysis may be due to limited variability in BMI among participants or the influence of other covariates such as glycemic and lipid control. However, these factors remain clinically important given their known associations implicated in the pathogenesis of diabetes progression and should continue to be monitored.

### Lifestyle and behavioral factors

Lifestyle factors, particularly high salt intake and physical inactivity, were significantly associated with complications. Patients with high salt consumption exhibited 57% increased odds of complications, suggesting that excessive sodium intake may exacerbate diabetes-related complications. High sodium intake is a known risk factor for hypertension, which in turn increases cardiovascular risks and accelerates the progression of vascular complications (57). Dietary interventions to reduce sodium intake to less than 2,400 mg per day have been shown to benefit T2DM patients and should be prioritized in diabetes management (58).

Physical inactivity was also significantly associated with higher odds of complications, with inactive patients exhibiting 1.75 times greater odds than active individuals. Regular exercise improves insulin sensitivity, glycemic control, and cardiovascular health, reinforcing its essential role in diabetes management (59). A prior meta-analysis reported a 29% reduction in cardiovascular disease incidence among active individuals, highlighting the inverse dose-response relationship between physical activity and diabetes complications (60). While organizations recommend 150 minutes of moderate aerobic activity per week (or its equivalent in vigorous activity), even modest increases in physical activity have been linked to reduced metabolic risk and lower mortality (61). These findings support the promotion and integration of structured exercise regimens as part of comprehensive diabetes care.

### Limitations of the study

This study has limitations that should be considered when interpreting the findings. First, its cross-sectional design limits causal inference between associated factors and diabetes complications. Second, self-reported data on lifestyle behaviors such as dietary salt intake, physical activity, alcohol use, foot care practice, and social support may be prone to recall and social desirability bias. Third, glycemic control was assessed using fasting blood sugar in some cases, which is less reliable than HbA1c for long-term monitoring. Fourth, excluding patients with incomplete records could introduce selection bias. Fifth, diagnostic inconsistencies may exist due to variability in clinician assessments and limited access to diagnostic tools. Sixth, as the study was conducted at a single specialized hospital, this may limit generalizability to a broader population. Lastly, missing data and unmeasured confounding may affect the findings.

## Conclusion

This study revealed a high burden of chronic complications among individuals with Type 2 diabetes mellitus in Southern Ethiopia, with nearly half affected and a considerable proportion having multimorbidity. Older age, female sex, longer diabetes duration, poor glycemic control, elevated triglycerides, high salt intake, and physical inactivity were factors significantly associated with chronic complications.

To address this problem, a comprehensive, multifaceted approach is needed. Routine screening, particularly for high-risk groups such as older adults and individuals with prolonged diabetes duration, should be strengthened. Patient-centered care should include individualized treatment plans that address sex-specific risks and improve access to diabetes education. Lifestyle modifications, such as dietary interventions to reduce sodium intake and structured physical activity programs, should be actively promoted. Additionally, regular monitoring and management of blood glucose, lipids, and blood pressure should be prioritized. National guidelines should incorporate preventive strategies, and healthcare institutions have to be strengthened to ensure timely diagnosis, effective management, and continuous follow-up care. Implementing these measures can significantly improve diabetes outcomes and reduce the burden of complications associated with T2DM.

## Data Availability

The datasets used and analyzed during the current study are available from the corresponding author upon reasonable request.

## Abbreviations

AOR: Adjusted Odds Ratio
BMI: Body Mass Index
COR: Crude Odds Ratio
DALYs: Disability-Adjusted Life Years
DM: Diabetes Mellitus
ESRD: End-Stage Renal Disease
IRB: Institutional Review Board
NCDs: Non-Communicable Diseases
SDGs: Sustainable Development Goals
T2DM: Type 2 Diabetes Mellitus
USD: United States Dollar
WHO: World Health Organization

## Availability of data and materials

The dataset includes sensitive participant information and cannot be publicly shared due to ethical restrictions approved by the Institutional Review Board (IRB) of Arba Minch University, and the terms of informed consent signed by study participants. Public disclosure would compromise participant privacy and confidentiality. Data access requests can be directed to the corresponding author and will require prior approval from the IRB.

## Author Contributions

BDA contributed to the study’s conception, design, data collection, and analysis, interpreted the results, and drafted the manuscript. YC and MBS contributed to the study’s conception, design, and interpretation of results and critically reviewed the manuscript. DHH, SMD, HEB, HW, and YH contributed to the interpretation of results and critically reviewed the manuscript. All authors have reviewed and approved the final version of the manuscript and agree to be accountable for all aspects of the work, ensuring the accuracy and integrity of the research.

## Acknowledgments

The authors wish to extend their heartfelt gratitude to Wolaita Sodo University Comprehensive Specialized Hospital for providing support and access to conduct this study. We are deeply thankful to all the patients who participated in this study. Special thanks also go to the dedicated data collectors for their hard work and commitment. This research would not have been possible without their invaluable contributions.

